# Favourable vaccine-induced SARS-CoV-2 specific T cell response profile in patients undergoing immune-modifying therapies

**DOI:** 10.1101/2022.02.21.22271127

**Authors:** Martin Qui, Nina Le Bert, Webber Pak Wo Chan, Malcolm Tan, Shou Kit Hang, Smrithi Hariharaputran, Jean Xiang Ying Sim, Jenny Guek Hong Low, Wei Ling Ng, Wei Yee Wan, Tiing Leong Ang, Antonio Bertoletti, Ennaliza Salazar

## Abstract

Patients undergoing immune-modifying therapies demonstrate a reduced humoral response after COVID-19 vaccination, but we lack a proper evaluation of the impact of such therapies on vaccine-induced T cell responses. Here, we longitudinally characterised humoral and Spike-specific T cell responses in inflammatory bowel disease (IBD) patients who are on antimetabolite therapy (azathioprine or methotrexate), TNF inhibitors and/or other biologic treatment (anti-integrin or anti-p40) after mRNA vaccination up to 3 months after completing two vaccine doses. We demonstrated that a Spike-specific T cell response is not only induced in treated IBD patients at levels similar to healthy individuals, but also sustained at higher magnitude, particularly in those treated with TNF inhibitor therapy. Furthermore, the Spike-specific T cell response in these patients is mainly preserved against mutations present in SARS-CoV-2 B.1.1.529 (Omicron) and characterized by a Th1/IL-10 cytokine profile. Thus, despite the humoral response defects, the favourable profile of vaccine-induced T cell responses might still provide a layer of COVID-19 protection to patients under immune-modifying therapies.

## INTRODUCTION

Immune-modifying agents are the treatment of choice for different chronic inflammatory diseases of autoimmune origin. Antimetabolites (azathioprine and methotrexate) or biologics, such as TNF inhibitors (adalimumab and infliximab), anti-p40 (ustekinumab) or anti-integrin (vedolizumab and etrolizumab) antibodies, are used alone or in combination to reduce inflammatory events in the gut (i.e. Crohn’s disease or ulcerative colitis), skin (i.e. psoriasis), joints (i.e. rheumatoid arthritis), or in multiple systems (i.e. systemic lupus erythematosus). While these agents reduce disease burden and improve quality of life (1), they are broadly considered immunosuppressive. The COVID-19 pandemic and the necessity to implement widespread vaccination sparked debate and research on the impact of these chronic therapies on the immunogenicity of SARS-CoV-2 vaccination (2–4).

Others have already shown that these therapies, particularly TNF inhibitors, reduce the ability of different COVID-19 vaccines (based on mRNA or Adenoviral vector) to produce Spike-specific antibodies (5–8) and recognize SARS-CoV-2 variants including B.1.617.2 (Delta) (4). Such results are expected, since reduced humoral responses to other vaccines (i.e. anti-pneumococcal, anti-HBV) was already demonstrated in patients similarly undergoing TNF inhibitor or other antimetabolite therapy (9–11), and since TNF-alpha has been demonstrated to play an important role in the coordinate maturation of humoral immunity (12).

Nevertheless antibodies can neither be considered the exclusive immunological parameter triggered by vaccination, nor the only determinant of its protective effect. Both mRNA- and adenoviral vector-based vaccines elicit humoral and cellular Spike-specific immunity (13–15) and an early induction of Spike-specific T cell responses associate with the early protective effect of mRNA vaccination (16). In addition, the apparent indispensability of coordinated humoral and cellular immune activation for rapid and successful control of SARS-CoV-2 infection (17), and the rise to global circulation of the Omicron variant (18) both highlight the importance of the vaccine-induced T cell response. While Spike mutations have conferred the Omicron variant with the ability to evade the majority of vaccine-induced neutralizing antibodies (19), Spike-specific T cell immunity remains mainly intact against the Omicron variant (20–23).

While these T cells might not play a role in preventing infection, their ability to recognize and lyse virus-infected cells likely represents an important antiviral mechanism that might prevent the unchecked spread of SARS-CoV-2 in the infected host (24). However, the impact that the different immune-modifying therapies exert on vaccine-induced Spike-specific cellular immunity has only started to be analysed (25), with initial evidence of preserved cellular immunity levels at least immediately after vaccination.

In this manuscript, we therefore studied a cohort of patients with inflammatory bowel diseases (IBD) who are antimetabolites (AM), TNF inhibitor (TNFi) and/or other biologic treatment (anti-integrin and anti-p40), and we characterized both cellular and humoral vaccine-induced Spike-specific immunity. Spike-specific immune responses were analysed from pre-vaccination, up until 3 months following the second dose of COVID-19 mRNA vaccination (BNT162b2 or mRNA-1273). Importantly, we designed an experimental plan to investigate not only the ability of vaccine to elicit “classical” Spike-specific T cell responses producing Th1 cytokines (IFN-γ and IL-2), but also the anti-inflammatory/regulatory IL-10 cytokine. The rationale of such an experimental design was based on data demonstrating that TNFi therapy mediates induction of IL-10 in T cells that likely contribute to its ability to dampen inflammation (26, 27).

The ability to modify the functional profile of classical Th1 T cells can be of particular importance in SARS-CoV-2 infection. The presence and induction of IL-10/IFN-γ producing SARS-CoV-2 specific T cells is associated with asymptomatic SARS-CoV-2 infection (28) and hybrid immunity (29), while their absence has been reported in severe COVID-19 (30). The induction of such T cells endowed with anti-inflammatory potential might be advantageous in the asymptomatic control of SARS-CoV-2 infection. Furthermore, due to the pervasion of the Omicron variant globally, the ability of vaccine-induced Spike-specific T cells to tolerate the amino acid mutations characteristic of the Omicron variant necessitates evaluation. We therefore tested directly ex vivo the impact that Omicron variant mutations exert on the Spike-specific T cells induced in IBD patients under different treatments.

## RESULTS

### Study population

During the study period, 83 patients had at least one blood sample analysed. Forty-three patients completed at least three visits while the rest were either lost to follow up or recruited after first or second vaccination. Of the 83 patients, 57 (69%) patients had Crohn’s disease and 26 (31%) had ulcerative colitis. Fifty three (64%) IBD patients and 18 (36%) HC were male (p=0.003), while median ages (years) in IBD (39) and HC group (40.5) were similar (p=0.65). Forty (48%) patients were on TNFi and 43 (52%) patients were on other non-TNFi immunotherapy. Baseline characteristics including duration of IBD diagnosis (years), disease phenotype and behaviour, and vaccine taken between TNFi and non-TNFi groups were similar as shown in Table 1. Among those undergoing TNF inhibitor therapy, 22 (55%) had an additional antimetabolite. Those on other non-TNFi consisted of 15 (35%) on an antimetabolite only, 7 (16%) on anti-p40 monotherapy and 9 (21%) on anti-integrin monotherapy. Those on an additional antimetabolite include 9 (21%) on anti-p40 (anti-IL-12/23) + AM and 3 (7%) on anti-integrin therapy + AM; those on anti-integrin + AM were excluded from analysis due to the lack of data points. Six patients were on concomitant steroid; 2 in the TNFi group and 4 in the non-TNFi group (p=0.74). Eight from the TNFi group and 6 patients from the non-TNFi group took the mRNA-1273 (Moderna) vaccine (p=0.66).

**Table 1.**
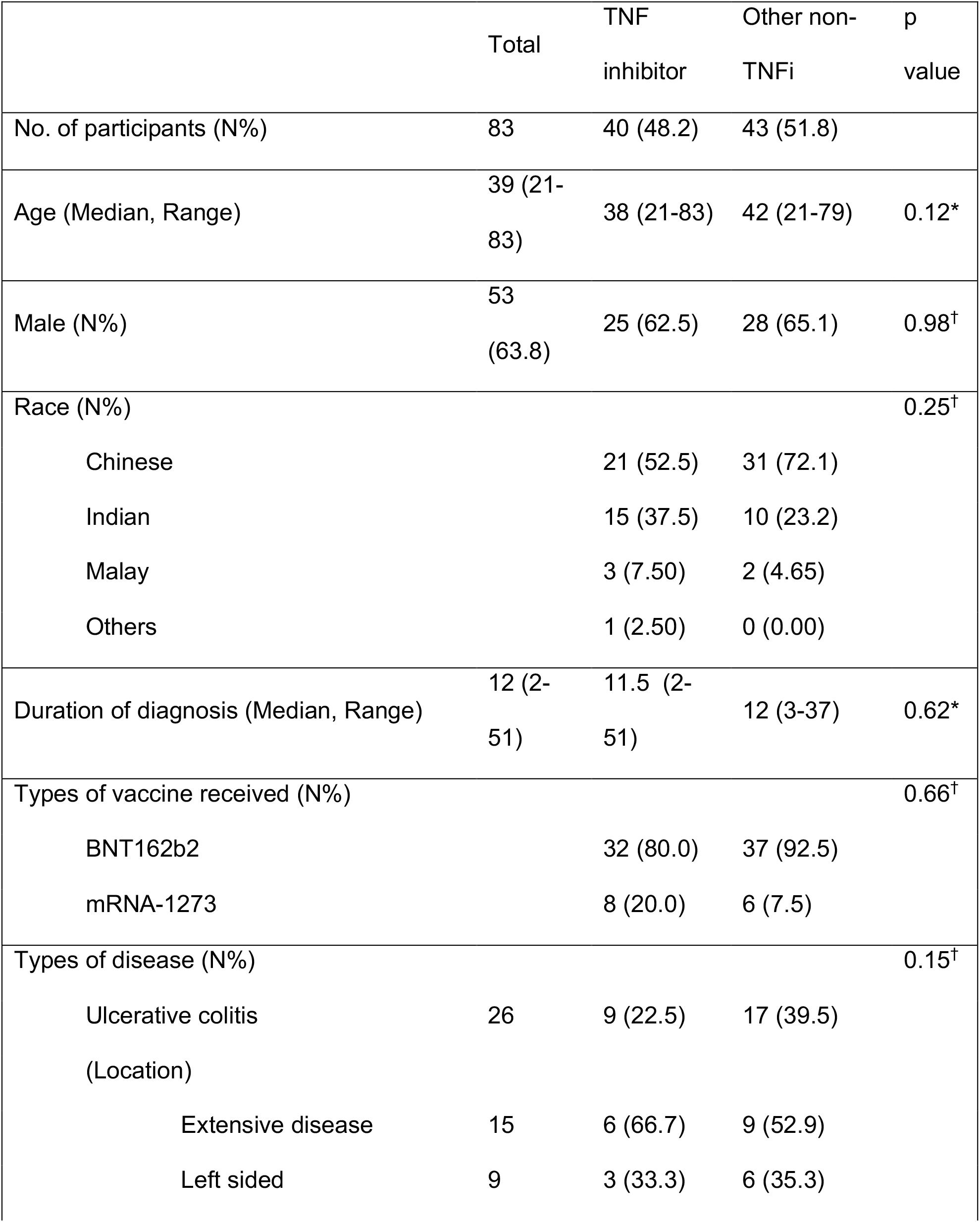
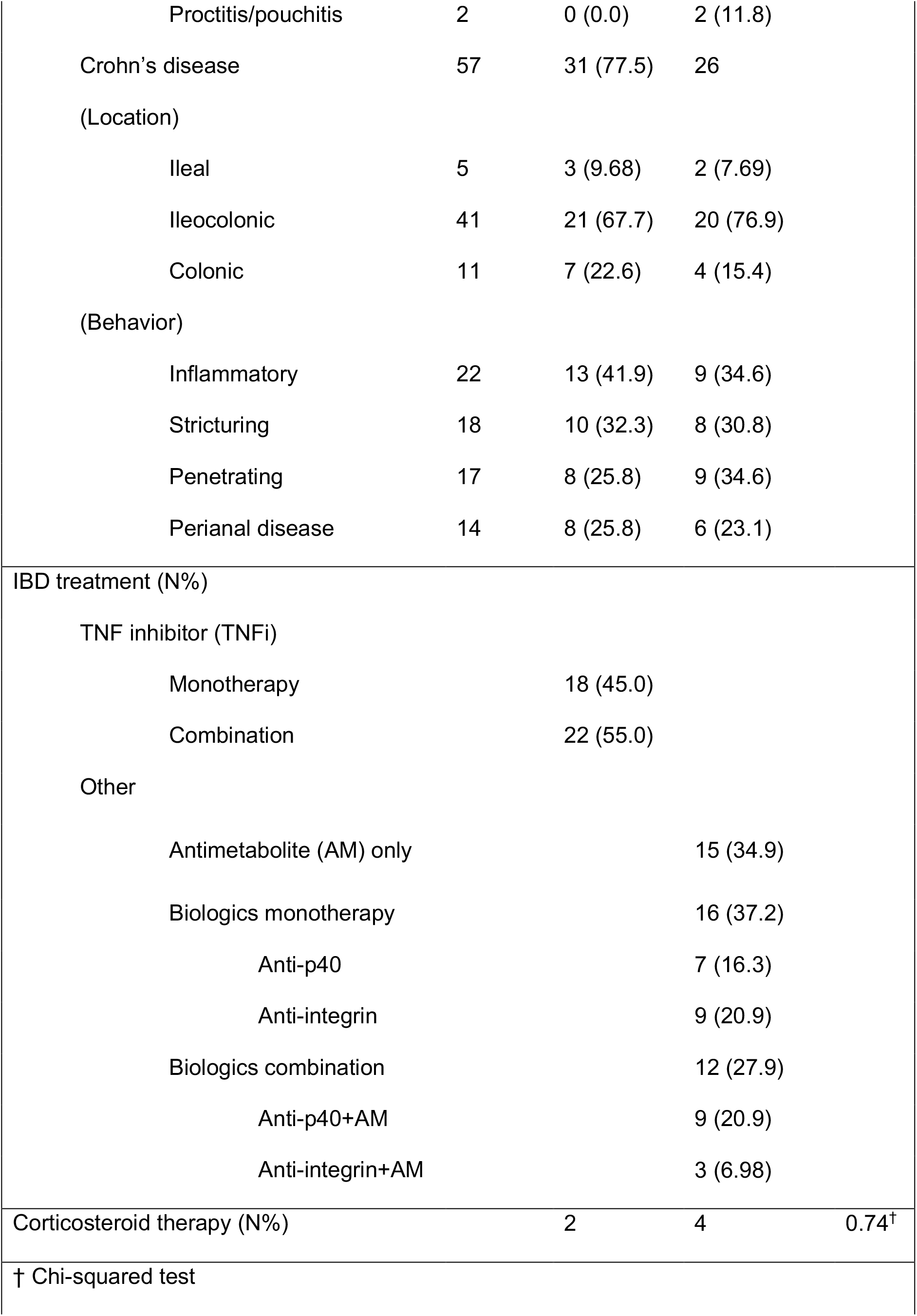
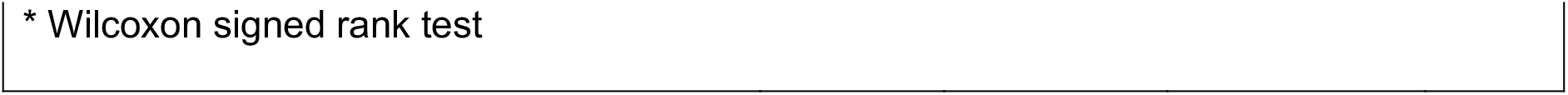
IBD patient demographics grouped according to TNF inhibitor status.

### Vaccine-induced humoral immunity

RBD IgG levels were quantified in response to COVID-19 mRNA vaccination both in the HCs and the IBD cohort (Figure 1A). In line with previous observations (5–7), at all post-vaccination timepoints D 21 (just before second mRNA dose), D 36 (two weeks fully vaccinated) and D 115 (three months fully vaccinated), the medians of IBD cohort humoral responses (geometric means/GMean of 176, 5545 and 949 AU/mL) were lower (p<0.005) than what observed in HC (GMean of 1212, 14951 and 2871 AU/mL) (Figure 1B).

**Figure 1.**
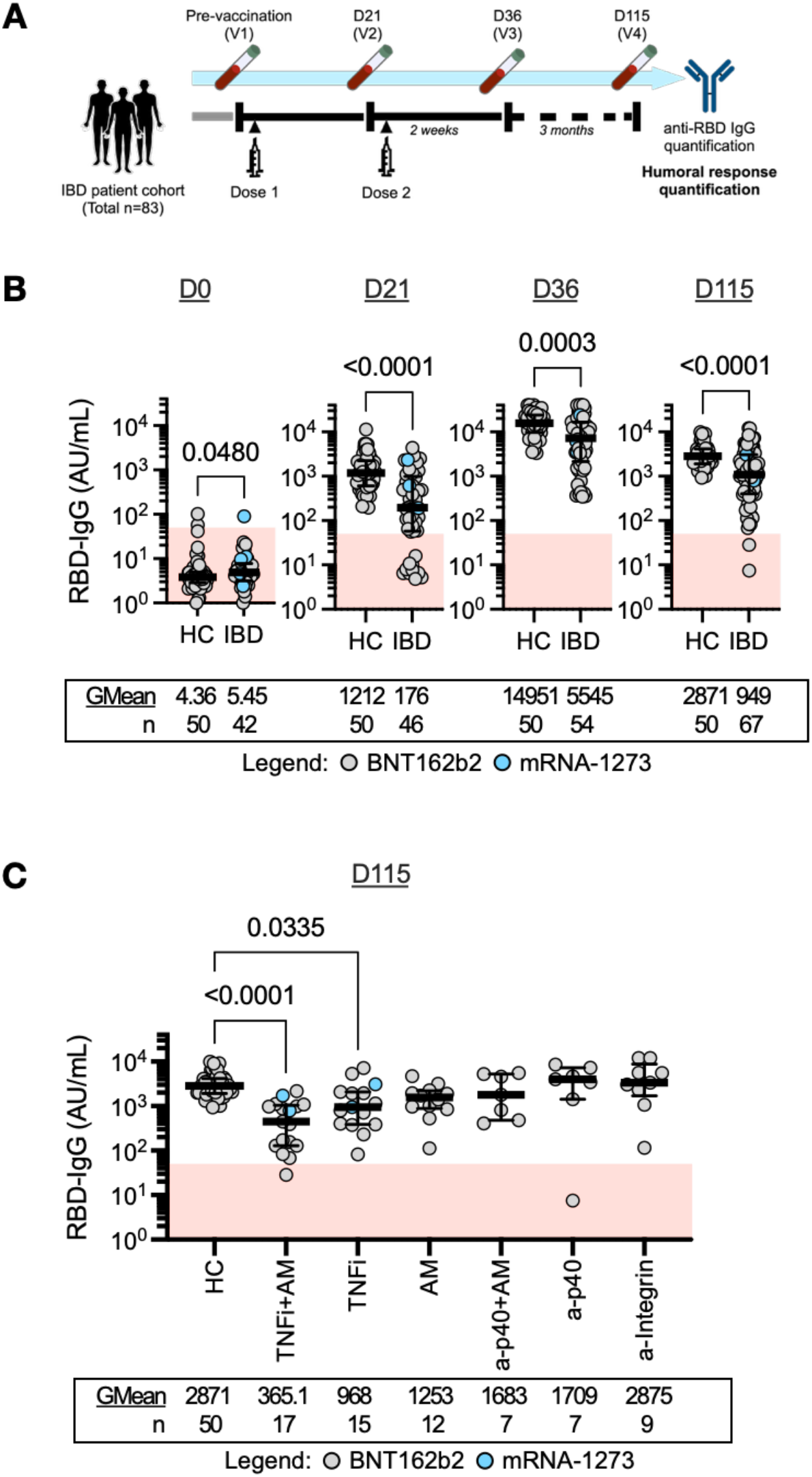
Humoral immunity is induced following COVID-19 mRNA vaccination. **(A)** Study design schematic for humoral response quantification. **(B)** Dot plots with median line (middle bar) and interquartile range (whiskers) of RBD IgG concentrations (AU/mL) from serum samples of the 2 study cohorts collected at different timepoints. Shaded red region denotes the area under the threshold for a positive test (50 AU/mL). Statistical analyses were performed by Wilcoxon signed rank test with p values indicated above the comparison line when significant (α=0.05). Geometric means (GMean) and number of data points (n) are indicated below each group. **(C)** Dot plots with median line (middle bar) and interquartile range (whiskers) of RBD IgG concentrations (AU/mL) from serum samples of HC and IBD patients grouped by treatment 3 months after completing their two-dose vaccination. Shaded red region denotes the area under the threshold for a positive test (50 AU/mL). Statistical analyses were performed by Kruskal-Wallis and Dunn’s test with p values shown above the comparison lines when significant (α=0.05). Geometric means (GMean) and number of data points (n) are indicated below each group

The deficiency of vaccine-induced anti-RBD antibodies was evident in patients undergoing therapy with TNFi monotherapy and even more in patients under combination with TNFi and an antimetabolite (TNFi+AM) (Figure 1C). No significant differences among anti-RBD titres 3 months after vaccination (D 115) in HC and IBD patients undergoing non-TNFi therapies were observed. Patients undergoing therapy with anti-p40 and anti-integrin (GMean 1709 and 2875 AU/mL) displayed anti-RBD titres that were indistinguishable from HC controls (GMean 2871 AU/mL) (Figure 1C).

### Vaccine-induced Spike-specific T cell responses

The magnitude and function of the Spike-specific T cell response induced by vaccination in HC and IBD patients was characterized directly in whole blood. A pool of 15-mer peptides covering the immunogenic regions of the SARS-CoV-2 S-protein (S pool) was used to measure Spike-specific T cell responses (Table S1). The quantity of Th1 cytokines (IFN-γ and IL-2) secreted in the plasma after peptide stimulation was quantified after overnight incubation (Figure 2A). This rapid quantitative assay has been demonstrated to possess identical sensitivities/specificities of conventional ELISpot assays (31).

**Figure 2.**
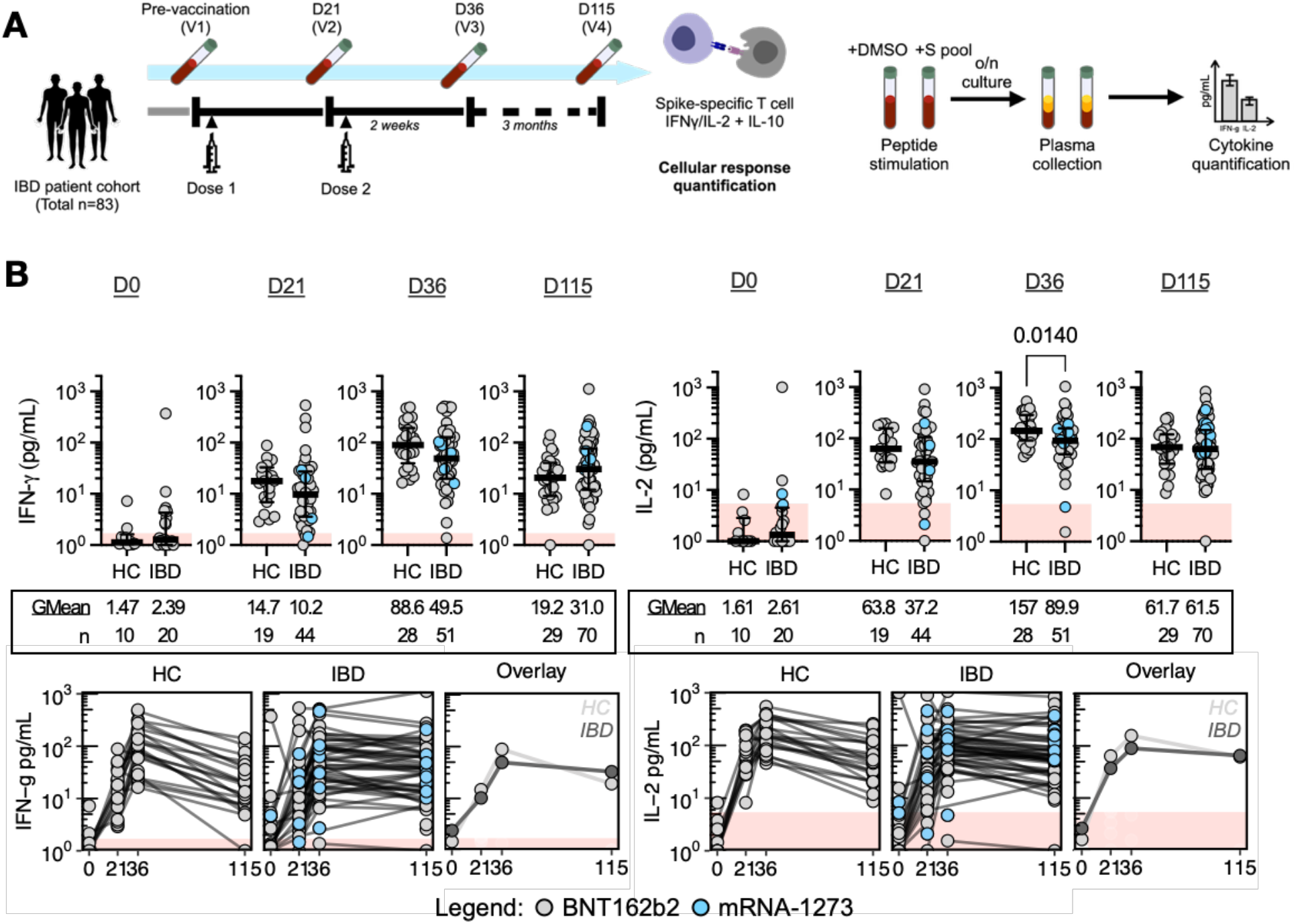
Cellular immunity is induced following COVID-19 mRNA vaccination. **(A)** Study design schematic for cellular response quantification (left panel) and overview of whole blood cytokine release assay for IFN-γ/IL-2 quantification. **(B)** Top panel: Dot plots with median line (middle bar) and interquartile range (whiskers) of IFN-γ (left) or IL-2 (right) concentrations (pg/mL) from S pool-stimulated whole blood supernatants of the 2 study cohorts collected at different timepoints. Shaded red region denote the area under the threshold for a positive test. Statistical analyses were performed by Wilcoxon signed rank test with p values indicated above the comparison line when significant (α=0.05). Geometric means (GMean) and number of data points (n) are indicated below each group. Bottom panel: Quantified IFN-γ or IL-2 concentrations (pg/mL) plotted against time, faceted by the two study cohorts. Shaded red region denote the area under the threshold for a positive test. Datapoints originating from the same participant are connected by gray lines. Data is summarized in the ‘Overlay’ plot with lines connecting the geometric means of each group at each sampling interval.

Before vaccination, whole blood supernatants of HC and IBD patients stimulated with S pool displayed median IFN-γ and IL-2 levels below threshold. Some whole blood supernatants from either cohort demonstrate cytokine production higher than unstimulated controls, consistent with the presence of Spike cross-reactive T cells already demonstrated in uninfected individuals (32, 33). Peptide-induced IFN-γ and/or IL-2 clearly increased in both HC and IBD patients after first (D 21) and second dose vaccination (D 36) in line with the previously demonstrated ability of mRNA vaccines to induce Spike-specific T cells (13). In particular, 2 weeks after second vaccination, all HC (28/28 for both IFN-γ and IL-2) and the majority of IBD patients under immune-modifying therapies possess a positive IFN-γ (50/51) and IL-2 (49/51) response (Figure 2B, top panel). Interestingly, while Spike peptide-induced cytokine levels decreased homogeneously in HC after 3 months (D 115), IBD patients demonstrated a more durable potential for cytokine induction (Figure 2B, bottom panel).

We then analysed cytokine levels induced in IBD patients under different treatment regimens. Notably, the durability of potential for cytokine induction is demonstrated broadly in the IBD cohort (Figure 3A). Furthermore, as shown in Figure 3B, patients treated with TNFi alone demonstrated not only more stable but also higher levels of IFN-γ and IL-2 responses than HC. Although few, we also noted that patients treated with anti-p40 biologics present high median levels of IL-2 production (n=7, GMean 229.6 pg/mL). Importantly, no specific treatment caused an inhibition of the quantity of IFN-γ and IL-2 3 months after the second vaccine dose (D 115) in comparison with HC; even the TNFi+AM group displaying lower Spike-specific humoral responses (Figure 1) demonstrated IFN-γ and IL-2 responses comparable to that in HC.

**Figure 3.**
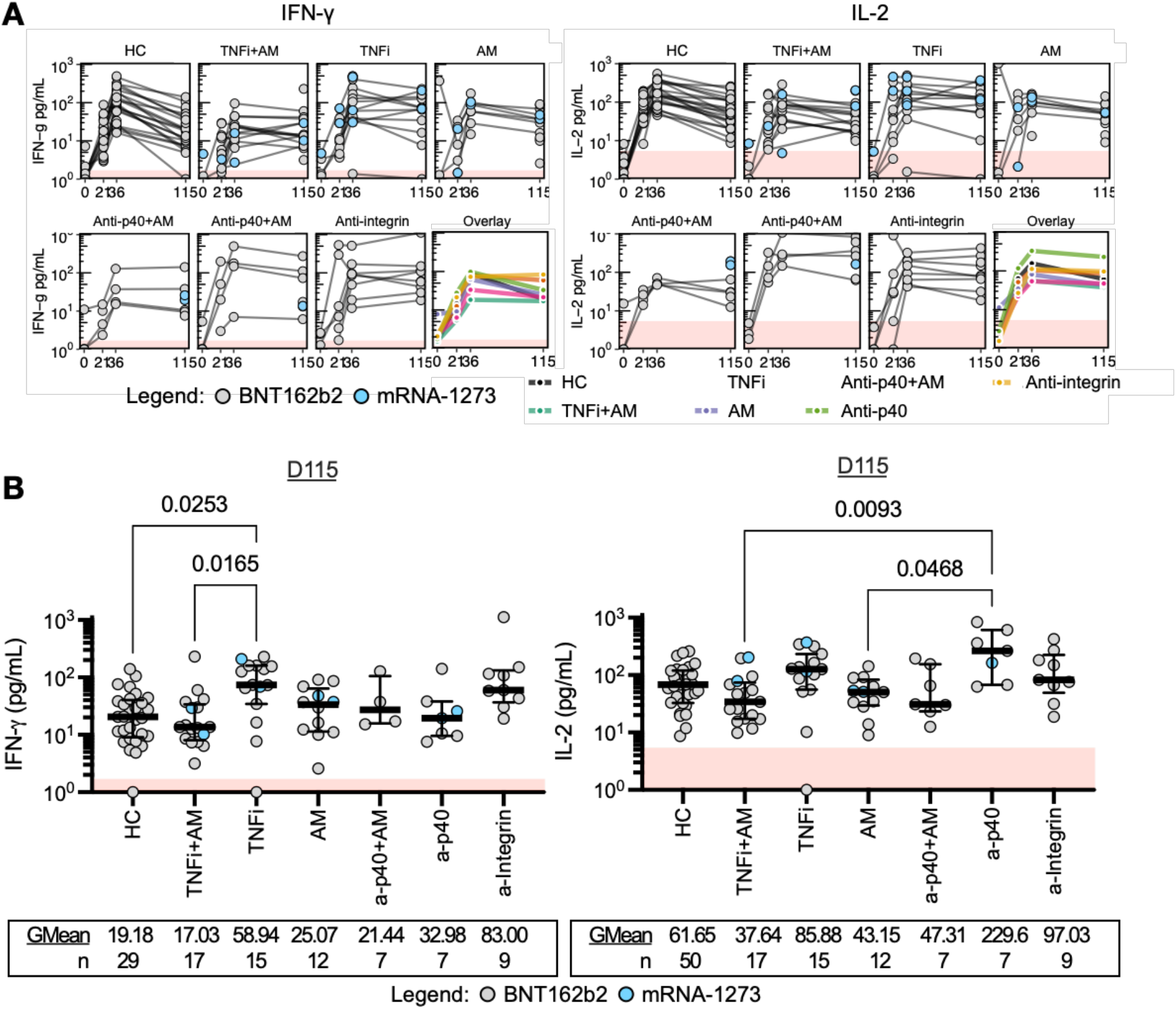
Durable T cell responses are demonstrated by patients under different immunotherapies. **(A)** Quantified IFN-γ or IL-2 concentrations (pg/mL) plotted against time of HC and IBD patients grouped by treatment. Shaded red region denote the area under the threshold for a positive test. Datapoints originating from the same participant are connected by gray lines. Data is summarized in the ‘Overlay’ plot with lines connecting the geometric means of each group at each sampling interval. **(B)** Dot plots with median line (middle bar) and interquartile range (whiskers) of quantified IFN-γ or IL-2 concentrations (pg/mL) from S pool-stimulated whole blood supernatants of HC and IBD patients grouped by treatment 3 months after completing their two-dose vaccination. Shaded red region denotes the area under the threshold for a positive test. Statistical analyses were performed by Kruskal-Wallis and Dunn’s test with p values shown above the comparison lines when significant (α=0.05). Geometric means (GMean) and number of data points (n) are indicated below each group.

We also compared the magnitude of Spike-specific T cell responses between IBD patients vaccinated with either BNT162b2 or mRNA-1273. No differences were observed in IFN-γ and IL-2 quantities at all time points, except an increased production of IL-2 in mRNA-1273 vaccinated IBD patients at D 115 (Figure S2).

Thus mRNA vaccination in IBD patients undergoing treatment with different immune-modifying therapies demonstrated a Spike-specific T cell cytokine responses that is not inferior to what is detectable in HC. Furthermore, TNFi therapy reduced the kinetics of contraction of T cell responses, resulting in a level of Spike-specific T cell responses 3 months after second vaccination (D 115) superior to that of HC.

### Spike-specific CD4+ and CD8+ T cells in vaccinated IBD patients and impact of Omicron variant mutations

To confirm that COVID-19 mRNA vaccination induces Spike-specific CD4+ and CD8+ T cells in IBD patients undergoing immune-modifying therapy, IBD patient PBMCs (n=7) collected at different time points were stimulated with Spike peptides pool and analysed for expression of activation markers on gated CD4+ and CD8+ T cells. Clear populations of CD4+ and CD8+ T cells were visualized and quantified upon peptide pool stimulation in 7 out of the 7 tested IBD patients (Figure 4A).

**Figure 4.**
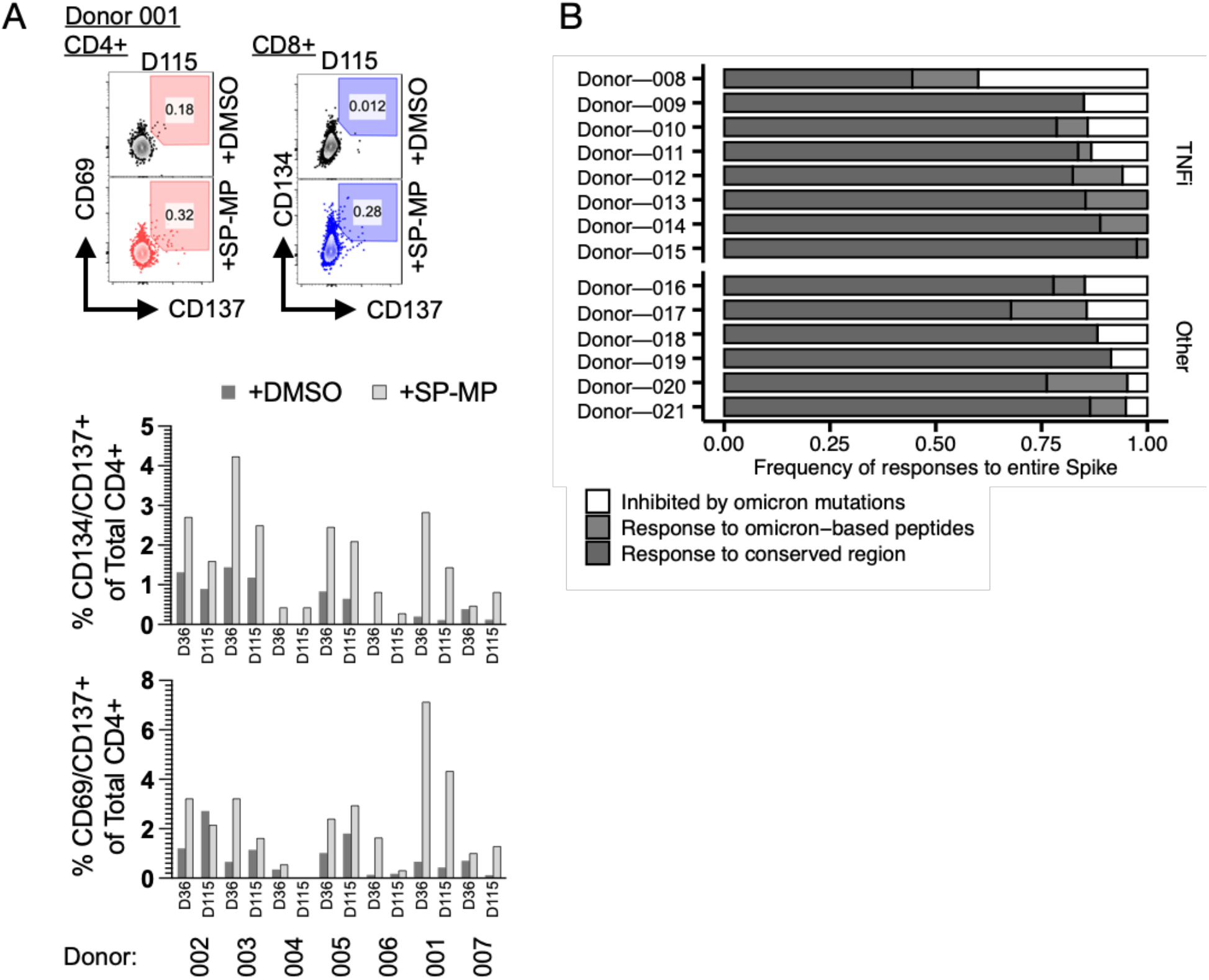
Spike-specific T cells are activated and function against ancestral or Omicron variant Spike. **(A)** Top: AIM assay to identify CD4+CD69+CD137+ (left, red) and CD8+CD134+CD137+ (right, blue) T cell populations with or without stimulation with Spike peptides. Activated cells are highlighted in the color-shaded region. Bottom: Summary frequencies of AIM+ cells identified from IBD patient PBMCs 2 weeks and 2 months after the second vaccine dose (n=7). **(B)** Bar graphs denoting component frequencies of the Spike-specific T cell response based on conserved and non-conserved regions between the ancestral and Omicron variant Spike. Spike-specific T cell response frequencies were derived from IFN-γ ELISpot of peptide-stimulated PBMCs (n=14).

We then analysed the impact of the mutations that characterize the Spike protein of the Omicron variant on the vaccine-induced Spike-specific T cells present in IBD patients (Figure 4B). Sample availability restricted our study to a subgroup of IBD patients (n=14). Patient PBMCs were stimulated with three peptide pools covering the entire Spike protein (253 peptides) of the ancestral SARS-CoV-2 (Table S2) and the regions mutated in the Omicron variant (67 peptides), with and without the amino acid-substitutions/deletions that characterize the SARS-CoV-2 Omicron variant (Table S3). We performed an IFN-γ ELISpot assay to quantify the frequency of SARS-CoV-2 specific T cells responding to conserved regions of the Spike protein, and to derive the frequency of responses altered by the variant-defining regions in the Omicron variant (Figure 4B). As already seen in healthy vaccinated individuals (20, 21), the Spike-specific T cell response to the Omicron variant is mainly preserved in the majority of patients irrespective of their treatment. Most Spike-specific T cells appear to target conserved regions, and an inhibition of more than 25% of the total Spike-specific T cell response due to Omicron mutations was observed in only 1 out of the 14 IBD patient sample tested.

### Immune-modifying therapies increase IL-10 production of Spike-specific T cells

Differences in the contraction kinetics of Spike peptide-induced IFN-γ and IL-2 detected in IBD patients undergoing TNFi therapy suggest that this treatment might modify vaccine-induced Spike-specific T cells. In addition, TNFi therapy has been shown to modify T cell function through expression of a transcriptional signature that upregulates IL-10 production in T cells (27). We therefore tested whether cytokine secretion profiles in whole blood supernatants after Spike-peptide stimulation contains not only classical Th1 cytokines IFN-γ and IL-2, but also IL-10. The quantity of IL-10 detected in IBD treated patients and HC before and after two dose vaccination was measured (Figure 5).

**Figure 5.**
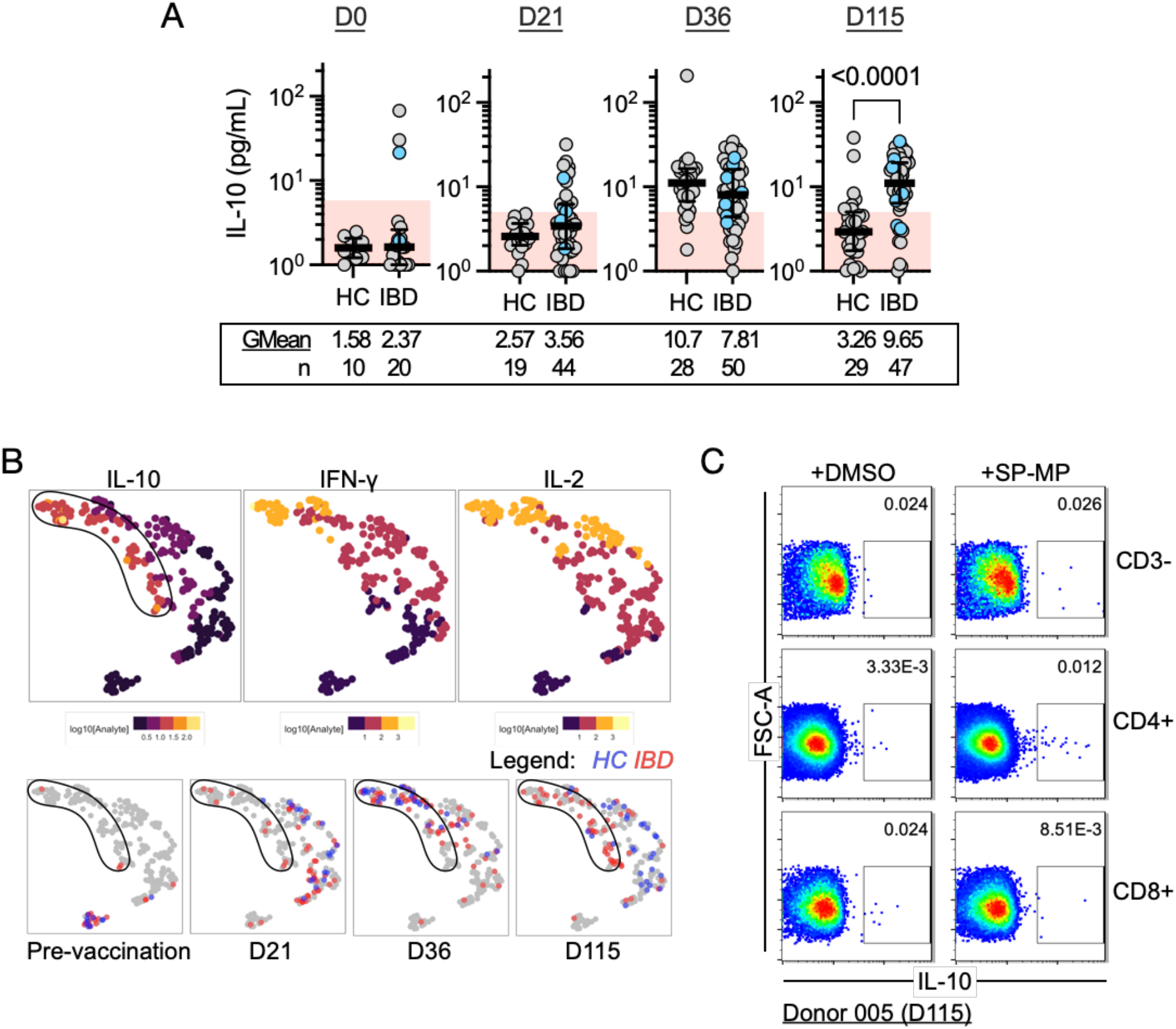
IL-10 delineates T cell cytokine response profile of individuals undergoing immune-modifying therapies. **(A)** Dot plots with median line (middle bar) and interquartile range (whiskers) of IL-10 concentrations (pg/mL) from S pool-stimulated whole blood supernatants of the 2 study cohorts collected at different timepoints. Shaded red region denotes the area under the threshold for a positive test. Statistical analyses were performed by Wilcoxon signed rank test with p values indicated above the comparison line when significant (α=0.05). Geometric means (GMean) and number of data points (n) are indicated below each group. **(B**) UMAP projections based on IL-10, IFN-γ and IL-2 quantities measured from each donor-timepoint. Top three panels display each point filled according to log10-transformed cytokine quantities (pg/mL). Bottom four panels display points filled according to study cohort at the respective timepoint. A shape is drawn enclosing a region mostly containing points with IL-10 values greater than 10 pg/mL. **(C)** Intracellular cytokine staining of donor PBMCs in CD3- or CD3+ (CD4+ and CD8+) compartments to identify IL-10+ cells with or without stimulation with Spike peptides

At both timepoints following first (D 21) and second dose vaccination (D 36), increased concentrations of IL-10 was detected in whole blood supernatants of HC and IBD patients relative to their respective pre-vaccination baselines. Furthermore, at 3 months after the second vaccine dose (D 115), no IL-10 could be detected in the majority of HC while we noticed a further increase in IL-10 induction in IBD patients (Figure 5A). This peak value (9.65 pg/mL), quantified at D 115, was low in comparison to corresponding IL-2 (61.5 pg/mL) and IFN-γ (31.5 pg/mL) responses in IBD patients.

To characterize the chronological evolution of cytokine profiles, we used UMAP to integrate quantified, log-transformed IL-10 data with IFN-γ and IL-2 for each donor-timepoint (Figure 5B). UMAP projections of datapoints originating from pre-vaccination samples of either HC or IBD patients formed a distinct cluster. Moreover, the datapoints co-segregated following first dose (D 21) and two weeks after second dose vaccination (D 36), further highlighting the similarities of cellular responses between the two cohorts. Notably, 3 months after completion of the two-dose regimen (D 115), the cytokine profiles diverge into distinct clusters, with IBD patient profiles coinciding with regions defined by higher IL-10, IFN-γ and IL-2.

To then confirm that Spike peptide pool stimulation induces IL-10 production in T cells, we performed direct ex vivo intracellular staining of T cells from TNFi treated patients (Figure 5C). Indeed, the low magnitude of cumulative IL-10 we observed in whole blood stimulated supernatants implied the identification of IL-10+ T cells to be a technically challenging feat, but a distinct population of IL-10+ cells was detected only in CD4+ T cells upon Spike peptide pool stimulation.

## DISCUSSION

The attenuated humoral responses detected in SARS-CoV-2 vaccinated patients under different immune-modifying treatments, particularly in those treated with TNFi therapy, have generally been interpreted to imply reduced vaccine immunogenicity, fuelling debate on the possible increased risk of severe COVID-19 in patients treated chronically with these immune-modifying therapies (2, 3, 6, 7). Here, by studying IBD patients under various regimens and vaccinated with the prevailing Spike-based mRNA vaccines, we demonstrated that a Spike-specific T cell response is not only induced in IBD treated patients to levels similar to that in HC, but persists longer and at higher levels in the ones treated with TNFi.

In contrast to the role of antibodies, T cells cannot prevent infection; instead, they excel in the clearance of intracellular pathogens either through recognition and lysis of infected cells or activating macrophages, and support B cell maturation *(32)*. Furthermore, since coordination between humoral and cellular arms of immunity is likely to be essential for rapid viral control and reduced pathogenicity (17), we cannot claim that the increased T cell immunogenicity directly translates into better protective efficacy of vaccination in patients under immune-modifying therapies. Nevertheless, these patients, particularly those undergoing TNFi therapy, are clearly able to mount a robust Spike-specific cellular immunity. Additionally, TNFi therapy does not abolish, but only reduces production of antibodies after mRNA vaccination. Previous observations and our own data show this. It is possible therefore to hypothesize that the presence of cellular immunity against Spike compensates for the observed humoral defect.

In this aspect, the demonstration provided here that vaccine-induced Spike-specific T cells of IBD patients are minimally altered on their ability to recognize Omicron variant Spike add a further layer of reassurance. Several recent works have shown that vaccine-induced Spike-specific T cells in healthy individuals is mainly preserved against the Omicron variant. Our data in IBD patients under different immune-modifying treatments demonstrate a similar pattern of reduced recognition only in a minority of tested patient samples. This finding suggests that, as in healthy vaccinated individuals (20, 21), the Spike-specific T cells of patients undergoing various immune-modifying regimens mount a multispecific T cell response against different conserved regions of Spike. Therefore, vaccinated patients undergoing TNFi therapy may develop reliable protection against severe disease. By analysing the kinetics of the Spike-specific T cell response, we observed that the higher levels of IFN-γ and IL-2 secretion present 3 months after vaccination in TNFi treated patients, in comparison to HC, does not derive from a higher level of vaccine-induced Spike-specific T cell induction at earlier time points, rather more likely from a propensity of the T cell response to persist longer. Such an observation might be explained by the ability of TNF-alpha to downregulate T cell expansion (34). Blocking TNF-alpha production can therefore cause a reduction of B cell maturation (12) but a progressive expansion of T cells (34).

The inhibition of TNF-alpha, directly through TNFi or indirectly through other immunomodulatory treatments, can also explain the simultaneous induction of IFN-γ, IL-2 and IL-10 found in IBD patients under different treatments. Blockade of the effect of TNF-alpha on T cells with TNF-alpha inhibitors is known to upregulate IL-10 in T cells (27). The presence of Spike-specific Th1 cells able to secrete IL-10 can be advantageous in SARS-CoV-2 infection. Animal models have shown that the ability of T cells to secrete IFN-γ and IL-10 simultaneously led to effective viral control without triggering severe pathological processes (35–37). Previously, we also observed that a pattern of cytokine production characterized by the simultaneous presence of IFN-γ, IL-2 and IL-10 constitute the T cell response detected in patients who control SARS-CoV-2 infection asymptomatically (28). The importance of IL-10/IFN-γ producing T cells have also been highlighted by two recent works: such a functional T cell profile was demonstrated to be defective in severe COVID-19 (30), while the presence of IL-10 producing Spike-specific T cells is characteristic of individuals with hybrid SARS-CoV-2 immunity (29) who demonstrate a robust immunity from re-infection (38, 39). Of note, the demonstration that mRNA vaccination in IBD patients undergoing TNF inhibition results in the induction of T cells with an IFN-γ/IL-2/IL-10 secretion profile suggests that similar functional profiles might likewise be induced in virus-specific T cells after SARS-CoV-2 infection, explaining the clinical evidence that SARS-CoV-2 infection in different patients undergoing TNFi treatment is generally mild (40–42). In our own study, 6 patients who eventually developed COVID-19 all had a mild disease course and did not require hospitalization; 4 of whom were on TNFi.

There are some limitations in this study: the most important one being that the bulk of T cell experiments were performed not by direct measurement of T cell quantity, but by measuring cytokines secreted in whole blood after specific peptide stimulation. However, we provide direct evidence orthogonally that Spike-specific CD4+ and CD8+ T cells were induced by vaccination in IBD patients and visualized IL-10+ T cells. Nevertheless, while this method does not directly quantify the number of T cells, it provides a standardised method well suited for longitudinal analysis of T cell responses in patients under different treatments. The simplicity of the assay reduces the inter-assay variability, and is directly performed on fresh whole blood, limiting the detrimental effects of freezing and thawing (43). Furthermore, since T cell functionality is analysed in whole blood, the immune-modifying therapies administered into the patients are present at therapeutic levels during the assay, mimicking, as we previously argued (44), more closely the situation in vivo.

In conclusion, we show here that mRNA vaccination in IBD patients under different immunomodulatory treatments triggers a robust cellular immune response amidst an attenuated humoral response. Particularly, patients under TNFi monotherapy demonstrate reduced kinetics of decline of Spike-specific T cell responses, and an ability to secrete a cytokine profile characterized by the simultaneous presence of IFN-γ, IL-2 and the anti-inflammatory IL-10 cytokine upon Spike encounter. Since this T cell functional profile has been preferentially associated with asymptomatic SARS-CoV-2 control, COVID-19 mRNA vaccination in individuals under such immunomodulatory therapies might still offer a layer of protection. Moreover, these may even offer some advantages in controlling SARS-CoV-2 infection with limited pathological sequelae.

## METHODS

### Study design

This is a prospective, observational study conducted to assess both humoral and cellular responses to mRNA-based COVID-19 vaccines (BNT162b2 and mRNA-1273) in inflammatory bowel disease (IBD) patients who were treated with antimetabolites, TNFi and/or other biologics from July 2021 to January 2022. Specifically, the included therapies were azathioprine or methotrexate for antimetabolites, adalimumab or infliximab for TNFi, ustekinumab for anti-p40, and vedolizumab or etrolizumab for anti-integrin. Patients have completed two same-dose vaccine courses three weeks apart with either one of the COVID-19 mRNA vaccines (n=83). The healthy control (HC) group included healthcare professionals not undergoing immune-modifying therapy (n=50). Patients younger than 18 years old, those with previous SARS-CoV-2 polymerase chain reaction-confirmed COVID-19, or pregnant women were all excluded. Samples were collected at baseline pre-vaccination (D 0), three weeks (D 21± 5 days) after first dose of vaccine, 2 weeks (D 36 ± 5 days) after second dose of vaccine and 3 months (D 115 ± 5 days) after second dose of vaccine. Due to the rapidity of vaccination uptake, recruitment of patients before vaccination became more challenging. Hence the protocol was extended to include IBD patients who were on antimetabolites/biologics and received their first and/or second dose of vaccine to be followed up longitudinally according to the aforementioned interval blood sampling.

### Quantification of humoral responses

Measurements were performed using the Abbott Architect i2000 automated analyser using the SARS-CoV-2 IgG II Quant assay, a chemiluminescent microparticle immunoassay (CMIA) for the quantitative detection of IgG targeting the receptor binding domain (RBD) of the S1 subunit of the spike protein of SARS-CoV-2. Results are expressed as AU/mL, where values ≥50.0 AU/mL are interpreted as positive.

### Quantification of cellular responses and analysis

We used a cytokine release assay (CRA) of whole peripheral blood stimulated using a SARS-CoV-2 spike-derived peptide (S) pool (Table S1) described previously (31). Freshly drawn whole blood (320µL; within 6 hours of venepuncture) was mixed with 80µL RPMI and stimulated with S pool peptides to a final peptide concentration of 2µg/mL or mixed with an equivalent amount of DMSO as control. Culture supernatants were collected 16 hours after culture and stored at −80°C until cytokine quantification. IFN-γ/IL-2 or IL-10 concentrations in plasma were quantified using an Ella machine (ProteinSimple) with microfluidic multiplex cartridges following the manufacturer’s instructions. Background cytokine levels quantified from DMSO controls were subtracted from the corresponding peptide pool stimulated samples. The threshold for a positive response was set at 10 times the lower limit of quantification for each cytokine (IFN-γ = 1.7 pg/mL; IL-2 = 5.4 pg/mL; IL-10 = 5.8 pg/mL). A pseudocount of 1 pg/mL was applied to the dataset for logistic transformation. Subsequently, log-transformed concentrations of each cytokine in all culture supernatants were projected onto UMAP space using 15 nearest neighbours (nn), min_dist of 0.2 and Euclidean distance.

### Peripheral blood mononuclear cell separation

Peripheral blood mononuclear cells (PBMC) from HBSS-diluted anticoagulated blood (1:1) were separated by Ficoll-Paque density gradient centrifugation. PBMCs were frozen in FBS containing 10% DMSO and stored in liquid nitrogen until use.

### ELISpot assay

ELISpot plates (Millipore) were coated with human IFN-γ antibody overnight at 4°C. Cryopreserved PBMCs were thawed and seeded at a density of 400,000 cells per well and stimulated with a respective peptide pool for 18 hours (2µg/mL) or an equivalent amount of DMSO (negative control). The plates were then incubated with human biotinylated IFN-γ detection antibody, followed by Streptavidin-AP and developed using the KPL BCIP/NBT Phosphatase Substrate. To quantify positive peptide-specific responses, twice the number of mean spots of the unstimulated wells were subtracted from the peptide-stimulated wells, and the results expressed as spot forming cells (SFU)/10^6^ PBMC. Results were excluded if negative control wells had >30 SFU/10^6^ PBMC or positive control wells (PMA/Ionomycin) were negative.

### Measurement of the impact of Omicron variant on total Spike-specific T cells

We directly tested donor PBMCs by IFN-γ ELISpot for reactivity against the ancestral or Omicron variant Spike protein. To quantify total responses to the ancestral Spike, we used a 10-amino acid overlapping 15-mer peptide pool (SP-MP) covering the entire Spike protein listed in Table S2 (1273 amino acids). For Omicron variant Spike responses, we designed two peptide pools (Table S3): one consisting of ancestral-derived Spike peptides covering the variable regions (termed the “Spike Hotspot-Ancestral” pool) and another consisting of the Omicron-derived Spike peptides covering the same region (termed the “Spike Hotspot-Omicron” pool). From this, we can derive the total spot forming units (SFU) formed against the entire Omicron variant Spike using the equation below:

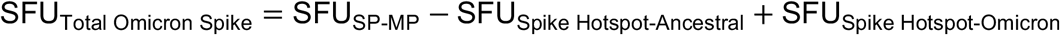

From this, % inhibition due to variation in Omicron variant Spike sequences may be quantified using the equation below:

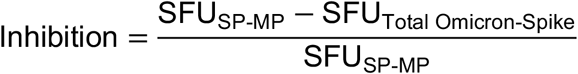

### Activation-induced marker (AIM) assay

For each condition, 1 million PBMCs in 150µL AIM-V + 2% AB were stimulated for 24 hr at 37°C with a megapool (2µg/mL) of 15-mer peptides encompassing the full spike protein (SP-MP) or an equivalent amount of DMSO in the presence of 1µg/mL CD28/CD49d co-stimulation (BD). Cells were then washed in FACS buffer with 2mM EDTA, then stained with surface markers anti-CD3-BV650, anti-CD4-AF700, anti-CD8-V500, anti-CD69-PE/Dazzle594, anti-CD134-PerCP-Cy5.5 and anti-CD-137-PE-Cy5 diluted in FACS buffer (RT for 25 minutes). Dead cells were excluded using the Fixable Yellow Live/Dead fixable cell stain kit (Invitrogen). After 2 more washes in FACS buffer, the cells were resuspended in PBS + 1% FA prior to analysis. The gating strategy is outlined in Figure S1, and the staining reagents used are outlined in Table S4.

### Intracellular cytokine staining

For each condition, 1 million PBMCs in 150µL AIM-V + 2% AB were stimulated for 24 hr at 37°C with SP-MP (2µg/mL) or an equivalent amount of DMSO in the presence of 1µg/mL CD28/CD49d co-stimulation (BD). In the last 4 hours, 1µg/mL Brefeldin A and 0.5X Monensin (Biolegend) were added. Cells were then washed in FACS buffer containing 2mM EDTA, then stained with surface markers anti-CD3-BV650, anti-CD4-AF700 and anti-CD8-V500, diluted in FACS buffer (RT for 25 minutes). Dead cells were excluded using the Fixable Yellow Live/Dead fixable cell stain kit (Invitrogen). Cells were washed twice in FACS buffer and fixed in Cytofix/Cytoperm (BD) for 20 minutes on ice. Cells were then washed with Perm/Wash (BD) solution prior to intracellular staining with anti-IL-10-BV421 diluted in Perm/Wash. After 2 more washes in FACS buffer, the cells were resuspended in PBS + 1% FA prior to analysis. The gating strategy is outlined in Figure S1, and the staining reagents used are outlined in Table S4.

### Flow cytometry

All flow cytometry samples were analysed using cryopreserved cells which were thawed and resuspended in AIM-V media supplemented with 2% AB serum. Samples were stained accordingly and fixed in PBS + 1% FA. Acquisition was performed on a BD-LSR II Analyzer (BD) within 24 hours and analysed with FlowJo software (BD)

### Statistics and data analysis

Statistical analyses were performed using R Statistical Software (version 4.0.3) (ggpubr::stat_compare_means) and GraphPad Prism 9. For analysis of the study population, Wilcoxon signed rank test and Chi-square tests were used as indicated. Median values in each group for humoral, cellular, IL-10 and T cell subset analysis were compared by Kruskal-Wallis test (with Dunn’s post-hoc multiple comparison test) or Wilcoxon signed rank test. Where applicable, statistical tests used and the definitions are indicated in the figure legends. P values <0.05 were considered statistically significant. Data from flow cytometry was analyzed using FlowJo software (BD).

### Study approval

The study protocol was reviewed and approved by Institutional Research Board of Singhealth (IRB no: 2021/2398). All donors provided written consent for enrolment.

## Data Availability

All data produced in the present work are contained in the manuscript

## AUTHOR CONTRIBUTIONS

The study was funded by the National Centre for Infectious Diseases Catalyst Grant and the National Research Fund Competitive Research Programme. MQ, NLB, AB and ES conceptualized and designed the experiments. WYW and WLN performed performed experiments for measuring humoral responses. MQ, HSK and SH performed experiments for measuring cellular responses. MQ, NLB, AB and ES analysed the data. MQ and ES prepared the figures and tables. SXYJ, JGHL and ES acquired funding for the project. WPWC, MT, ES, JGHL and TLA collected donor samples. Writing was prepared and edited by MQ, NLB, WYW, AB and ES.

## ACKNOWLEDGEMENTS

We would like to acknowledge the contribution of SGH-IBD team members Dr Thomson Lim Chong Teik (MD), Dr Tay Shu Wen (MD), Loy Kia Lan (Nurse practitioner), Lim Teong Guan (Pharmacist), Dr Ong Wan Chee (Pharmacist), Dr Valerie Ng Yun Ting (Pharmacist) and Amanda Wong Shi Yi (Pharmacist) who helped in recruiting patients. We also would like to thank Abigail Yeong, Tan Hui Fang and Stephanie Ren for coordinating the logistics of the study and follow-ups for all the study participants. Lastly we would like to thank all our patients who took their time off to participate in the study. This study was supported by funds under the NCID Catalyst Grant and administered by the National Centre for Infectious Diseases. Any opinions, findings and conclusions or recommendations expressed in this material are those of the author(s) and do not reflect the views of NCID.

