## Supplementary material for "Favourable vaccine-induced SARS-CoV-2 specific T cell response profile in patients undergoing immune-modifying therapies": Sup tables and figures

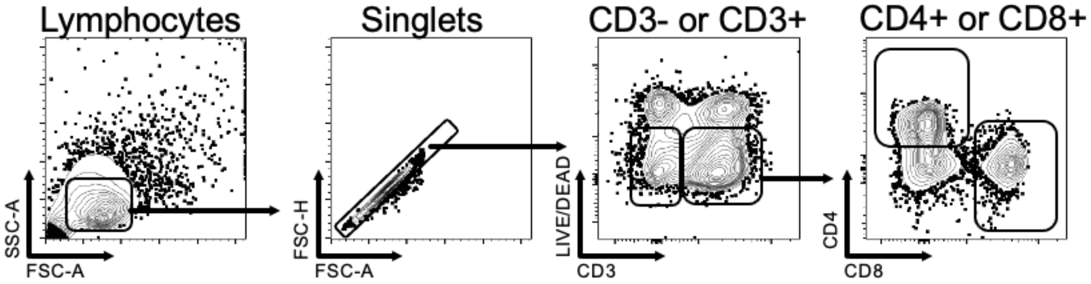


**Figure S1. Gating strategy of T cell subsets for activation-induced markers assay/intracellular cytokine staining.** Bivariate contour plots with outliers displaying a measured parameter obtained from flow cytometry indicated at the lower left corner of each plot. Gates are outlined in each plot

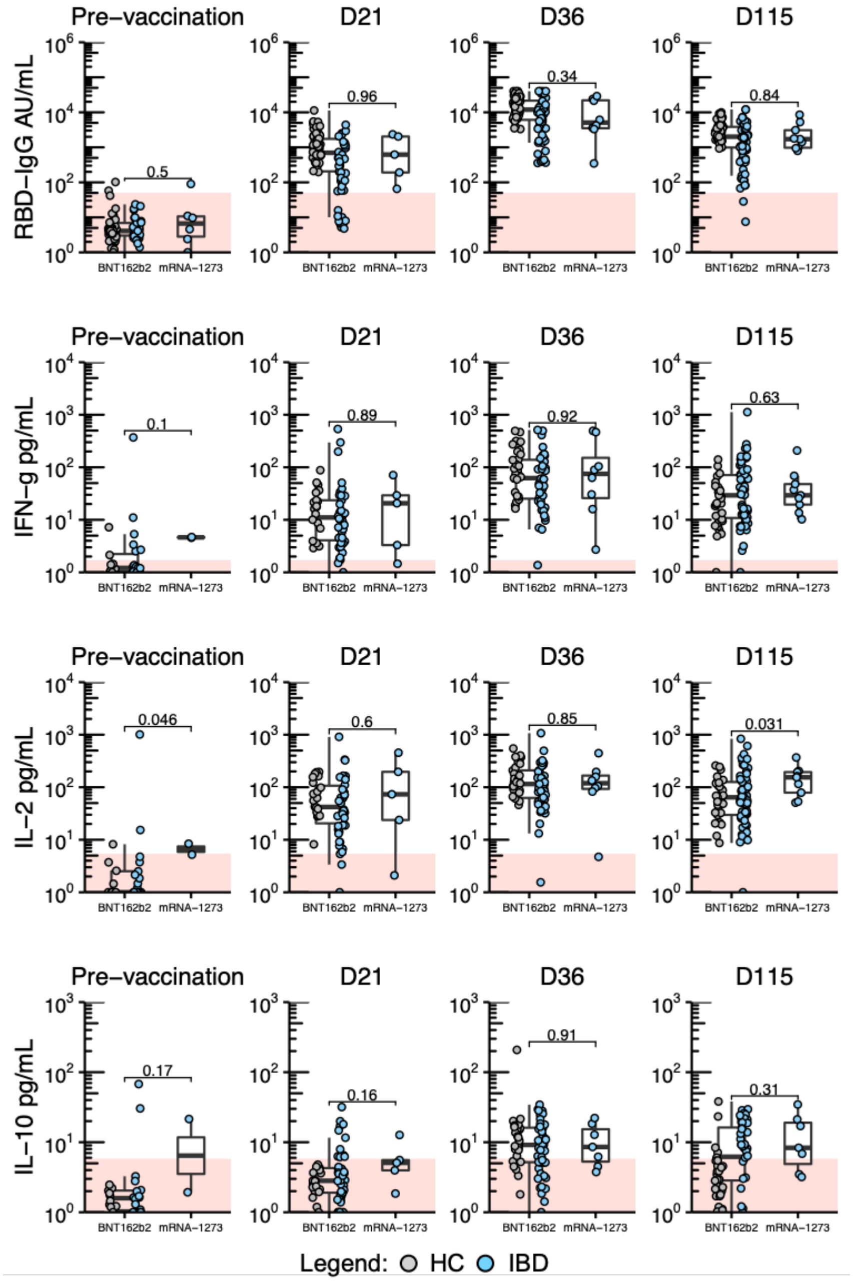


**Figure S2. Responses based on mRNA vaccine administered faceted by study timepoint.** Box and whisker plots with median line. Shaded red region indicates the threshold for a positive test depending on the analyte (RBD-IgG: 50 AU/mL; IFN-g: 1.7 pg/mL; IL-2: 5.4 pg/mL; IL-10: 5.8 pg/mL). Legend is indicated at the bottom. Statistical analyses were performed by MWW tests with p values indicated above the comparison line.

Table S1. SARS-CoV-2 S pool peptide library

| Peptide # | Peptide Sequence | aa |
|---|---|---|
| **1** | IRGWIFGTTLDSKTQ | 101-115 |
| **2** | FGTTLDSKTQSLLIV | 106-120 |
| **3** | CTFEYVSQPFLMDLE | 166-180 |
| **4** | VSQPFLMDLEGKQGN | 171-185 |
| **5** | TRFQTLLALHRSYLT | 236-250 |
| **6** | LLALHRSYLTPGDSS | 241-255 |
| **7** | RSYLTPGDSSSGWTA | 246-260 |
| **8** | CALDPLSETKCTLKS | 291-305 |
| **9** | LSETKCTLKSFTVEK | 296-310 |
| **10** | CTLKSFTVEKGIYQT | 301-315 |
| **11** | FTVEKGIYQTSNFRV | 306-320 |
| **12** | GIYQTSNFRVQPTES | 311-325 |
| **13** | SASFSTFKCYGVSPT | 371-385 |
| **14** | TFKCYGVSPTKLNDL | 376-390 |
| **15** | YNYKLPDDFTGCVIA | 421-435 |
| **16** | WNSNNLDSKVGGNYN | 436-450 |
| **17** | LDSKVGGNYNYLYRL | 441-455 |
| **18** | GGNYNYLYRLFRKSN | 446-460 |
| **19** | YLYRLFRKSNLKPFE | 451-465 |
| **20** | FRKSNLKPFERDIST | 456-470 |
| **21** | LKPFERDISTEIYQA | 461-475 |
| **22** | GPKKSTNLVKNKCVN | 526-540 |
| **23** | TNLVKNKCVNFNFNG | 531-545 |
| **24** | FNFNGLTGTGVLTES | 541-555 |
| **25** | LTGTGVLTESNKKFL | 546-560 |
| **26** | RAGCLIGAEHVNNSY | 646-660 |
| **27** | IGAEHVNNSYECDIP | 651-665 |
| **28** | SVASQSIIAYTMSLG | 686-700 |
| **29** | SIIAYTMSLGAENSV | 691-705 |
| **30** | TMSLGAENSVAYSNN | 696-710 |
| **31** | STECSNLLLQYGSFC | 746-760 |
| **32** | NLLLQYGSFCTQLNR | 751-765 |
| **33** | KNTQEVFAQVKQIYK | 776-790 |
| **34** | VFAQVKQIYKTPPIK | 781-795 |

| Peptide # | Peptide Sequence | aa |
|---|---|---|
| **35** | KQIYKTPPIKDFGGF | 786-800 |
| **36** | TPPIKDFGGFNFSQI | 791-805 |
| **37** | NFSQILPDPSKPSKR | 801-815 |
| **38** | AGFIKQYGDCLGDIA | 831-845 |
| **39** | QYGDCLGDIAARDLI | 836-850 |
| **40** | GAALQIPFAMQMAYR | 891-905 |
| **41** | QMAYRFNGIGVTQNV | 901-915 |
| **42** | FNGIGVTQNVLYENQ | 906-920 |
| **43** | DSLSSTASALGKLQD | 936-950 |
| **44** | TASALGKLQDVVNQN | 941-955 |
| **45** | AQALNTLVKQLSSNF | 956-970 |
| **46** | VLNDILSRLDKVEAE | 976-990 |
| **47** | LITGRLQSLQTYVTQ | 996-1010 |
| **48** | QLIRAAEIRASANLA | 1011-1025 |
| **49** | AEIRASANLAATKMS | 1016-1030 |
| **50** | APHGVVFLHVTYVPA | 1056-1070 |
| **51** | HWFVTQRNFYEPQII | 1101-1115 |
| **52** | KEIDRLNEVAKNLNE | 1181-1195 |
| **53** | LNEVAKNLNESLIDL | 1186-1200 |
| **54** | KNLNESLIDLQELGK | 1191-1205 |
| **55** | IWLGFIAGLIAIVMV | 1216-1230 |

Table S2: SARS-CoV-2 SP-MP pool peptide library

| Peptide # | Peptide Sequence | aa |
|---|---|---|
| **1** | MFVFLVLLPLVSSQC | 1-15 |
| **2** | VLLPLVSSQCVNLTT | 6-20 |
| **3** | VSSQCVNLTTRTQLP | 11-25 |
| **4** | VNLTTRTQLPPAYTN | 16-30 |
| **5** | RTQLPPAYTNSFTRG | 21-35 |
| **6** | PAYTNSFTRGVYYPD | 26-40 |
| **7** | SFTRGVYYPDKVFRS | 31-45 |
| **8** | VYYPDKVFRSSVLHS | 36-50 |
| **9** | KVFRSSVLHSTQDLF | 41-55 |
| **10** | SVLHSTQDLFLPFFS | 46-60 |
| **11** | TQDLFLPFFSNVTWF | 51-65 |
| **12** | LPFFSNVTWFHAIHV | 56-70 |
| **13** | NVTWFHAIHVSGTNG | 61-75 |
| **14** | HAIHVSGTNGTKRFD | 66-80 |
| **15** | SGTNGTKRFDNPVLP | 71-85 |
| **16** | TKRFDNPVLPFNDGV | 76-90 |
| **17** | NPVLPFNDGVYFAST | 81-95 |
| **18** | FNDGVYFASTEKSNI | 86-100 |
| **19** | YFASTEKSNIIRGWI | 91-105 |
| **20** | EKSNIIRGWIFGTTL | 96-110 |
| **21** | IRGWIFGTTLDSKTQ | 101-115 |
| **22** | FGTTLDSKTQSLLIV | 106-120 |
| **23** | DSKTQSLLIVNNATN | 111-125 |
| **24** | SLLIVNNATNVVIKV | 116-130 |
| **25** | NNATNVVIKVCEFQF | 121-135 |
| **26** | VVIKVCEFQFCNDPF | 126-140 |
| **27** | CEFQFCNDPFLGVYY | 131-145 |
| **28** | CNDPFLGVYYHKNNK | 136-150 |
| **29** | LGVYYHKNNKSWMES | 141-155 |
| **30** | HKNNKSWMESEFRVY | 146-160 |
| **31** | SWMESEFRVYSSANN | 151-165 |
| **32** | EFRVYSSANNCTFEY | 156-170 |
| **33** | SSANNCTFEYVSQPF | 161-175 |
| **34** | CTFEYVSQPFLMDLE | 166-180 |

| Peptide # | Peptide Sequence | aa |
|---|---|---|
| **35** | VSQPFLMDLEGKQGN | 171-185 |
| **36** | LMDLEGKQGNFKNLR | 176-190 |
| **37** | GKQGNFKNLREFVFK | 181-195 |
| **38** | FKNLREFVFKNIDGY | 186-200 |
| **39** | EFVFKNIDGYFKIYS | 191-205 |
| **40** | NIDGYFKIYSKHTPI | 196-210 |
| **41** | FKIYSKHTPINLVRD | 201-215 |
| **42** | KHTPINLVRDLPQGF | 206-220 |
| **43** | NLVRDLPQGFSALEP | 211-225 |
| **44** | LPQGFSALEPLVDLP | 216-230 |
| **45** | SALEPLVDLPIGINI | 221-235 |
| **46** | LVDLPIGINITRFQT | 226-240 |
| **47** | IGINITRFQTLLALH | 231-245 |
| **48** | TRFQTLLALHRSYLT | 236-250 |
| **49** | LLALHRSYLTPGDSS | 241-255 |
| **50** | RSYLTPGDSSSGWTA | 246-260 |
| **51** | PGDSSSGWTAGAAAY | 251-265 |
| **52** | SGWTAGAAAYYVGYL | 256-270 |
| **53** | GAAAYYVGYLQPRTF | 261-275 |
| **54** | YVGYLQPRTFLLKYN | 266-280 |
| **55** | QPRTFLLKYNENGTI | 271-285 |
| **56** | LLKYNENGTITDAVD | 276-290 |
| **57** | ENGTITDAVDCALDP | 281-295 |
| **58** | TDAVDCALDPLSETK | 286-300 |
| **59** | CALDPLSETKCTLKS | 291-305 |
| **60** | LSETKCTLKSFTVEK | 296-310 |
| **61** | CTLKSFTVEKGIYQT | 301-315 |
| **62** | FTVEKGIYQTSNFRV | 306-320 |
| **63** | GIYQTSNFRVQPTES | 311-325 |
| **64** | SNFRVQPTESIVRFP | 316-330 |
| **65** | QPTESIVRFPNITNL | 321-335 |
| **66** | IVRFPNITNLCPFGE | 326-340 |
| **67** | NITNLCPFGEVFNAT | 331-345 |
| **68** | CPFGEVFNATRFASV | 336-350 |
| **69** | VFNATRFASVYAWNR | 341-355 |

| Peptide # | Peptide Sequence | aa |
|---|---|---|
| **70** | RFASVYAWNRKRISN | 346-360 |
| **71** | YAWNRKRISNCVADY | 351-365 |
| **72** | KRISNCVADYSVLYN | 356-370 |
| **73** | CVADYSVLYNSASFS | 361-375 |
| **74** | SVLYNSASFSTFKCY | 366-380 |
| **75** | SASFSTFKCYGVSPT | 371-385 |
| **76** | TFKCYGVSPTKLNDL | 376-390 |
| **77** | GVSPTKLNDLCFTNV | 381-395 |
| **78** | KLNDLCFTNVYADSF | 386-400 |
| **79** | CFTNVYADSFVIRGD | 391-405 |
| **80** | YADSFVIRGDEVRQI | 396-410 |
| **81** | VIRGDEVRQIAPGQT | 401-415 |
| **82** | EVRQIAPGQTGKIAD | 406-420 |
| **83** | APGQTGKIADYNYKL | 411-425 |
| **84** | GKIADYNYKLPDDFT | 416-430 |
| **85** | YNYKLPDDFTGCVIA | 421-435 |
| **86** | PDDFTGCVIAWNSNN | 426-440 |
| **87** | GCVIAWNSNNLDSKV | 431-445 |
| **88** | WNSNNLDSKVGGNYN | 436-450 |
| **89** | LDSKVGGNYNYLYRL | 441-455 |
| **90** | GGNYNYLYRLFRKSN | 446-460 |
| **91** | YLYRLFRKSNLKPFE | 451-465 |
| **92** | FRKSNLKPFERDIST | 456-470 |
| **93** | LKPFERDISTEIYQA | 461-475 |
| **94** | RDISTEIYQAGSTPC | 466-480 |
| **95** | EIYQAGSTPCNGVEG | 471-485 |
| **96** | GSTPCNGVEGFNCYF | 476-490 |
| **97** | NGVEGFNCYFPLQSY | 481-495 |
| **98** | FNCYFPLQSYGFQPT | 486-500 |
| **99** | PLQSYGFQPTNGVGY | 491-505 |
| **100** | GFQPTNGVGYQPYRV | 496-510 |
| **101** | NGVGYQPYRVVVLSF | 501-515 |
| **102** | QPYRVVVLSFELLHA | 506-520 |
| **103** | VVLSFELLHAPATVC | 511-525 |

| Peptide # | Peptide Sequence | aa |
|---|---|---|
| **104** | ELLHAPATVCGPKKS | 516-530 |
| **105** | PATVCGPKKSTNLVK | 521-535 |
| **106** | GPKKSTNLVKNKCVN | 526-540 |
| **107** | TNLVKNKCVNFNFNG | 531-545 |
| **108** | NKCVNFNFNGLTGTG | 536-550 |
| **109** | FNFNGLTGTGVLTES | 541-555 |
| **110** | LTGTGVLTESNKKFL | 546-560 |
| **111** | VLTESNKKFLPFQQF | 551-565 |
| **112** | NKKFLPFQQFGRDIA | 556-570 |
| **113** | PFQQFGRDIADTTDA | 561-575 |
| **114** | GRDIADTTDAVRDPQ | 566-580 |
| **115** | DTTDAVRDPQTLEIL | 571-585 |
| **116** | VRDPQTLEILDITPC | 576-590 |
| **117** | TLEILDITPCSFGGV | 581-595 |
| **118** | DITPCSFGGVSVITP | 586-600 |
| **119** | SFGGVSVITPGTNTS | 591-605 |
| **120** | SVITPGTNTSNQVAV | 596-610 |
| **121** | GTNTSNQVAVLYQDV | 601-615 |
| **122** | NQVAVLYQDVNCTEV | 606-620 |
| **123** | LYQDVNCTEVPVAIH | 611-625 |
| **124** | NCTEVPVAIHADQLT | 616-630 |
| **125** | PVAIHADQLTPTWRV | 621-635 |
| **126** | ADQLTPTWRVYSTGS | 626-640 |
| **127** | PTWRVYSTGSNVFQT | 631-645 |
| **128** | YSTGSNVFQTRAGCL | 636-650 |
| **129** | NVFQTRAGCLIGAEH | 641-655 |
| **130** | RAGCLIGAEHVNNSY | 646-660 |
| **131** | IGAEHVNNSYECDIP | 651-665 |
| **132** | VNNSYECDIPIGAGI | 656-670 |
| **133** | ECDIPIGAGICASYQ | 661-675 |
| **134** | IGAGICASYQTQTNS | 666-680 |
| **135** | CASYQTQTNSPRRAR | 671-685 |
| **136** | TQTNSPRRARSVASQ | 676-690 |
| **137** | PRRARSVASQSIIAY | 681-695 |

| Peptide # | Peptide Sequence | aa |
|---|---|---|
| **138** | SVASQSIIAYTMSLG | 686-700 |
| **139** | SIIAYTMSLGAENSV | 691-705 |
| **140** | TMSLGAENSVAYSNN | 696-710 |
| **141** | AENSVAYSNNSIAIP | 701-715 |
| **142** | AYSNNSIAIPTNFTI | 706-720 |
| **143** | SIAIPTNFTISVTTE | 711-725 |
| **144** | TNFTISVTTEILPVS | 716-730 |
| **145** | SVTTEILPVSMTKTS | 721-735 |
| **146** | ILPVSMTKTSVDCTM | 726-740 |
| **147** | MTKTSVDCTMYICGD | 731-745 |
| **148** | VDCTMYICGDSTECS | 736-750 |
| **149** | YICGDSTECSNLLLQ | 741-755 |
| **150** | STECSNLLLQYGSFC | 746-760 |
| **151** | NLLLQYGSFCTQLNR | 751-765 |
| **152** | YGSFCTQLNRALTGI | 756-770 |
| **153** | TQLNRALTGIAVEQD | 761-775 |
| **154** | ALTGIAVEQDKNTQE | 766-780 |
| **155** | AVEQDKNTQEVFAQV | 771-785 |
| **156** | KNTQEVFAQVKQIYK | 776-790 |
| **157** | VFAQVKQIYKTPPIK | 781-795 |
| **158** | KQIYKTPPIKDFGGF | 786-800 |
| **159** | TPPIKDFGGFNFSQI | 791-805 |
| **160** | DFGGFNFSQILPDPS | 796-810 |
| **161** | NFSQILPDPSKPSKR | 801-815 |
| **162** | LPDPSKPSKRSFIED | 806-820 |
| **163** | KPSKRSFIEDLLFNK | 811-825 |
| **164** | SFIEDLLFNKVTLAD | 816-830 |
| **165** | LLFNKVTLADAGFIK | 821-835 |
| **166** | VTLADAGFIKQYGDC | 826-840 |
| **167** | AGFIKQYGDCLGDIA | 831-845 |
| **168** | QYGDCLGDIAARDLI | 836-850 |
| **169** | LGDIAARDLICAQKF | 841-855 |
| **170** | ARDLICAQKFNGLTV | 846-860 |
| **171** | CAQKFNGLTVLPPLL | 851-865 |

| Peptide # | Peptide Sequence | aa |
|---|---|---|
| **172** | NGLTVLPPLLTDEMI | 856-870 |
| **173** | LPPLLTDEMIAQYTS | 861-875 |
| **174** | TDEMIAQYTSALLAG | 866-880 |
| **175** | AQYTSALLAGTITSG | 871-885 |
| **176** | ALLAGTITSGWTFGA | 876-890 |
| **177** | TITSGWTFGAGAALQ | 881-895 |
| **178** | WTFGAGAALQIPFAM | 886-900 |
| **179** | GAALQIPFAMQMAYR | 891-905 |
| **180** | IPFAMQMAYRFNGIG | 896-910 |
| **181** | QMAYRFNGIGVTQNV | 901-915 |
| **182** | FNGIGVTQNVLYENQ | 906-920 |
| **183** | VTQNVLYENQKLIAN | 911-925 |
| **184** | LYENQKLIANQFNSA | 916-930 |
| **185** | KLIANQFNSAIGKIQ | 921-935 |
| **186** | QFNSAIGKIQDSLSS | 926-940 |
| **187** | IGKIQDSLSSTASAL | 931-945 |
| **188** | DSLSSTASALGKLQD | 936-950 |
| **189** | TASALGKLQDVVNQN | 941-955 |
| **190** | GKLQDVVNQNAQALN | 946-960 |
| **191** | VVNQNAQALNTLVKQ | 951-965 |
| **192** | AQALNTLVKQLSSNF | 956-970 |
| **193** | TLVKQLSSNFGAISS | 961-975 |
| **194** | LSSNFGAISSVLNDI | 966-980 |
| **195** | GAISSVLNDILSRLD | 971-985 |
| **196** | VLNDILSRLDKVEAE | 976-990 |
| **197** | LSRLDKVEAEVQIDR | 981-995 |
| **198** | KVEAEVQIDRLITGR | 986-1000 |
| **199** | VQIDRLITGRLQSLQ | 991-1005 |
| **200** | LITGRLQSLQTYVTQ | 996-1010 |
| **201** | LQSLQTYVTQQLIRA | 1001-1015 |
| **202** | TYVTQQLIRAAEIRA | 1006-1020 |
| **203** | QLIRAAEIRASANLA | 1011-1025 |
| **204** | AEIRASANLAATKMS | 1016-1030 |
| **205** | SANLAATKMSECVLG | 1021-1035 |

| Peptide # | Peptide Sequence | aa |
|---|---|---|
| **206** | ATKMSECVLGQSKRV | 1026-1040 |
| **207** | ECVLGQSKRVDFCGK | 1031-1045 |
| **208** | QSKRVDFCGKGYHLM | 1036-1050 |
| **209** | DFCGKGYHLMSFPQS | 1041-1055 |
| **210** | GYHLMSFPQSAPHGV | 1046-1060 |
| **211** | SFPQSAPHGVVFLHV | 1051-1065 |
| **212** | APHGVVFLHVTYVPA | 1056-1070 |
| **213** | VFLHVTYVPAQEKNF | 1061-1075 |
| **214** | TYVPAQEKNFTTAPA | 1066-1080 |
| **215** | QEKNFTTAPAICHDG | 1071-1085 |
| **216** | TTAPAICHDGKAHFP | 1076-1090 |
| **217** | ICHDGKAHFPREGVF | 1081-1095 |
| **218** | KAHFPREGVFVSNGT | 1086-1100 |
| **219** | REGVFVSNGTHWFVT | 1091-1105 |
| **220** | VSNGTHWFVTQRNFY | 1096-1110 |
| **221** | HWFVTQRNFYEPQII | 1101-1115 |
| **222** | QRNFYEPQIITTDNT | 1106-1120 |
| **223** | EPQIITTDNTFVSGN | 1111-1125 |
| **224** | TTDNTFVSGNCDVVI | 1116-1130 |
| **225** | FVSGNCDVVIGIVNN | 1121-1135 |
| **226** | CDVVIGIVNNTVYDP | 1126-1140 |
| **227** | GIVNNTVYDPLQPEL | 1131-1145 |
| **228** | TVYDPLQPELDSFKE | 1136-1150 |
| **229** | LQPELDSFKEELDKY | 1141-1155 |
| **230** | DSFKEELDKYFKNHT | 1146-1160 |
| **231** | ELDKYFKNHTSPDVD | 1151-1165 |
| **232** | FKNHTSPDVDLGDIS | 1156-1170 |
| **233** | SPDVDLGDISGINAS | 1161-1175 |
| **234** | LGDISGINASVVNIQ | 1166-1180 |
| **235** | GINASVVNIQKEIDR | 1171-1185 |
| **236** | VVNIQKEIDRLNEVA | 1176-1190 |
| **237** | KEIDRLNEVAKNLNE | 1181-1195 |
| **238** | LNEVAKNLNESLIDL | 1186-1200 |
| **239** | KNLNESLIDLQELGK | 1191-1205 |

| Peptide # | Peptide Sequence | aa |
|---|---|---|
| **240** | SLIDLQELGKYEQYI | 1196-1210 |
| **241** | QELGKYEQYIKWPWY | 1201-1215 |
| **242** | YEQYIKWPWYIWLGF | 1206-1220 |
| **243** | KWPWYIWLGFIAGLI | 1211-1225 |
| **244** | IWLGFIAGLIAIVMV | 1216-1230 |
| **245** | IAGLIAIVMVTIMLC | 1221-1235 |
| **246** | AIVMVTIMLCCMTSC | 1226-1240 |
| **247** | TIMLCCMTSCCSCLK | 1231-1245 |
| **248** | CMTSCCSCLKGCCSC | 1236-1250 |
| **249** | CSCLKGCCSCGSCCK | 1241-1255 |
| **250** | GCCSCGSCCKFDEDD | 1246-1260 |
| **251** | GSCCKFDEDDSEPVL | 1251-1265 |
| **252** | FDEDDSEPVLKGVKL | 1256-1270 |
| **253** | SEPVLKGVKLHYT | 1261-1273 |

Table S3. SARS-CoV-2 Spike Hotspot-Ancestral and Hotspot-Omicron peptide libraries used to study responses to omicron

| Peptide # | Spike Hotspot-Ancestral | Spike Hotspot-Omicron | aa |
|---|---|---|---|
| **1** | LPFFSNVTWFHAIHV | LPFFSNVTWFHVISG | 56-70 |
| **2** | NVTWFHAIHVSGTNG | NVTWFHVISGTNGTK | 61-75 |
| **3** | HAIHVSGTNGTKRFD | HVISGTNGTKRFDNP | 66-80 |
| **4** | FNDGVYFASTEKSNI | FNDGVYFASIEKSNI | 86-100 |
| **5** | YFASTEKSNIIRGWI | YFASIEKSNIIRGWI | 91-105 |
| **6** | CEFQFCNDPFLGVYY | CEFQFCNDPFLDHKN | 131-145 |
| **7** | CNDPFLGVYYHKNNK | CNDPFLDHKNNKSWM | 136-150 |
| **8** | LGVYYHKNNKSWMES | LDHKNNKSWMESEFR | 141-155 |
| **9** | FKIYSKHTPINLVRD | FKIYSKHTPIIVEPE | 201-215 |
| **10** | KHTPINLVRDLPQGF | KHTPIIVEPERDLPQ | 206-220 |
| **11** | NLVRDLPQGFSALEP | IVEPERDLPQGFSAL | 211-225 |
| **12** | IVRFPNITNLCPFGE | IVRFPNITNLCPFDE | 326-340 |
| **13** | NITNLCPFGEVFNAT | NITNLCPFDEVFNAT | 331-345 |
| **14** | CPFGEVFNATRFASV | CPFDEVFNATRFASV | 336-350 |
| **15** | CVADYSVLYNSASFS | CVADYSVLYNLAPFF | 361-375 |
| **16** | SVLYNSASFSTFKCY | SVLYNLAPFFTFKCY | 366-380 |
| **17** | SASFSTFKCYGVSPT | LAPFFTFKCYGVSPT | 371-385 |
| **18** | EVRQIAPGQTGKIAD | EVRQIAPGQTGNIAD | 406-420 |
| **19** | APGQTGKIADYNYKL | APGQTGNIADYNYKL | 411-425 |
| **20** | GKIADYNYKLPDDFT | GNIADYNYKLPDDFT | 416-430 |
| **21** | PDDFTGCVIAWNSNN | PDDFTGCVIAWNSNK | 426-440 |
| **22** | GCVIAWNSNNLDSKV | GCVIAWNSNKLDSKV | 431-445 |
| **23** | WNSNNLDSKVGGNYN | WNSNKLDSKVSGNYN | 436-450 |
| **24** | LDSKVGGNYNYLYRL | LDSKVSGNYNYLYRL | 441-455 |
| **25** | GGNYNYLYRLFRKSN | SGNYNYLYRLFRKSN | 446-460 |
| **26** | RDISTEIYQAGSTPC | RDISTEIYQAGNKPC | 466-480 |
| **27** | EIYQAGSTPCNGVEG | EIYQAGNKPCNGVAG | 471-485 |
| **28** | GSTPCNGVEGFNCYF | GNKPCNGVAGFNCYF | 476-490 |
| **29** | NGVEGFNCYFPLQSY | NGVAGFNCYFPLRSY | 481-495 |
| **30** | FNCYFPLQSYGFQPT | FNCYFPLRSYSFRPT | 486-500 |
| **31** | PLQSYGFQPTNGVGY | PLRSYSFRPTYGVGH | 491-505 |
| **32** | GFQPTNGVGYQPYRV | SFRPTYGVGHQPYRV | 496-510 |
| **33** | NGVGYQPYRVVVLSF | YGVGHQPYRVVVLSF | 501-515 |
| **34** | NKCVNFNFNGLTGTG | NKCVNFNFNGLKGTG | 536-550 |
| **35** | FNFNGLTGTGVLTES | FNFNGLKGTGVLTES | 541-555 |
| **36** | LTGTGVLTESNKKFL | LKGTGVLTESNKKFL | 546-560 |

| Peptide # | Spike Hotspot-Ancestral | Spike Hotspot-Omicron | aa |
|---|---|---|---|
| **37** | GTNTSNQVAVLYQDV | GTNTSNQVAVLYQGV | 601-615 |
| **38** | NQVAVLYQDVNCTEV | NQVAVLYQGVNCTEV | 606-620 |
| **39** | LYQDVNCTEVPVAIH | LYQGVNCTEVPVAIH | 611-625 |
| **40** | NVFQTRAGCLIGAEH | NVFQTRAGCLIGAEY | 641-655 |
| **41** | RAGCLIGAEHVNNSY | RAGCLIGAEYVNNSY | 646-660 |
| **42** | IGAEHVNNSYECDIP | IGAEYVNNSYECDIP | 651-665 |
| **43** | IGAGICASYQTQTNS | IGAGICASYQTQTKS | 666-680 |
| **44** | CASYQTQTNSPRRAR | CASYQTQTKSHRRAR | 671-685 |
| **45** | TQTNSPRRARSVASQ | TQTKSHRRARSVASQ | 676-690 |
| **46** | PRRARSVASQSIIAY | HRRARSVASQSIIAY | 681-695 |
| **47** | SIIAYTMSLGAENSV | SIIAYTMSLGVENSV | 691-705 |
| **48** | TMSLGAENSVAYSNN | TMSLGVENSVAYSNN | 696-710 |
| **49** | AENSVAYSNNSIAIP | VENSVAYSNNSIAIP | 701-715 |
| **50** | NLLLQYGSFCTQLNR | NLLLQYGSFCTQLKR | 751-765 |
| **51** | YGSFCTQLNRALTGI | YGSFCTQLKRALTGI | 756-770 |
| **52** | TQLNRALTGIAVEQD | TQLKRALTGIAVEQD | 761-775 |
| **53** | KQIYKTPPIKDFGGF | KQIYKTPPIKYFGGF | 786-800 |
| **54** | TPPIKDFGGFNFSQI | TPPIKYFGGFNFSQI | 791-805 |
| **55** | DFGGFNFSQILPDPS | YFGGFNFSQILPDPS | 796-810 |
| **56** | ARDLICAQKFNGLTV | ARDLICAQKFKGLTV | 846-860 |
| **57** | CAQKFNGLTVLPPLL | CAQKFKGLTVLPPLL | 851-865 |
| **58** | NGLTVLPPLLTDEMI | KGLTVLPPLLTDEMI | 856-870 |
| **59** | TASALGKLQDVVNQN | TASALGKLQDVVNHN | 941-955 |
| **60** | GKLQDVVNQNAQALN | GKLQDVVNHNAQALN | 946-960 |
| **61** | VVNQNAQALNTLVKQ | VVNHNAQALNTLVKQ | 951-965 |
| **62** | AQALNTLVKQLSSNF | AQALNTLVKQLSSKF | 956-970 |
| **63** | TLVKQLSSNFGAISS | TLVKQLSSKFGAISS | 961-975 |
| **64** | LSSNFGAISSVLNDI | LSSKFGAISSVLNDI | 966-980 |
| **65** | GAISSVLNDILSRLD | GAISSVLNDIFSRLD | 971-985 |
| **66** | VLNDILSRLDKVEAE | VLNDIFSRLDKVEAE | 976-990 |
| **67** | LSRLDKVEAEVQIDR | FSRLDKVEAEVQIDR | 981-995 |

Table S4. List of labelling reagents used for flow cytometry

| Target | Color | Clone | Brand | RRID/Cat. no | Dilution (in 100µL) |
|---|---|---|---|---|---|
| Anti-human CD3 | BV650 | SP34-2 | BD Horizon™ | AB_2738486 | 2.5 |
| Anti-human CD4 | AF700 | RPA-T4 | Biolegend | AB_493743 | 2.5 |
| Anti-human CD8 | V500 | RPA-T8 | BD Horizon™ | AB_1937333 | 0.5 |
| Anti-human IL-10 | BV421 | JES3-9D7 | Biolegend | AB_10896947 | 6 |
| Anti-human CD69 | PE/Dazzle594 | FN50 | Biolegend | AB_2564276 | 2 |
| Anti-human CD134 | PerCP-Cy5.5 | Ber-ACT35 | Biolegend | AB_10720986 | 2 |
| Anti-human CD137 | PE-Cy5 | 4B4-1 | BD Horizon™ | AB_394067 | 5 |
| LIVE/DEAD™ Fixable Yellow Dead Cell Stain Kit (405nm excitation) | | | ThermoFisher | Cat. L34959 | 0.1 |
